# Transcriptome and DNA methylome analysis of peripheral blood samples reveals incomplete restoration and transposable element activation after 3-month recovery of COVID-19

**DOI:** 10.1101/2022.04.19.22274029

**Authors:** Ying Yin, Xiao-zhao Liu, Qing Tian, Yi-xian Fan, Zhen Ye, Tian-qing Meng, Gong-hong Wei, Cheng-liang Xiong, Honggang Li, Ximiao He, Li-quan Zhou

## Abstract

Comprehensive analyses showed that SARS-CoV-2 infection caused COVID-19 and induced strong immune responses and sometimes severe illnesses. However, cellular features of recovered patients and long-term health consequences remain largely unexplored. In this study, we collected peripheral blood samples from recovered COVID-19 patients (average age of 35.7 years old) from Hubei province, China, 3 months after discharge; and carried out RNA-seq and whole-genome bisulfite sequencing (WGBS) to identify hallmarks of recovered COVID-19 patients. Our analyses showed significant changes both in expression and DNA methylation of genes and transposable elements (TEs) in recovered COVID-19 patients. We identified 639 misregulated genes and 18516 differentially methylated regions (DMRs) in total. Genes with aberrant expression and DMRs were found to be associated with immune responses and other related biological processes, implicating prolonged overreaction of the immune system in response to SARS-CoV-2 infection. Notably, a significant amount of TEs were aberrantly activated and TE activation was positively correlated with COVID-19 severity. Moreover, differentially methylated TEs may regulate adjacent gene expression as regulatory elements. Those identified transcriptomic and epigenomic signatures define and drive the features of recovered COVID-19 patients, helping determine the risks of long COVID-19, and providing guidance for clinical intervention.

## INTRODUCTION

Emerging SARS-CoV-2 coronavirus which causes coronavirus disease 2019 (COVID-19) and results in complicated health issues has expanded rapidly and swept the whole world, threatening global public health(Guan et al., 2020; Huang et al., 2020). As of November 2021, the COVID-19 pandemic has resulted in approximate 260 million confirmed cases, including 5 million deaths worldwide(Dashboard, 2021; Shen et al., 2014). Identified key receptors for SARS-CoV-2 infection include ACE2 (Zhou et al., 2020), TMPRSS2(Hoffmann et al., 2020) and NRP1(Cantuti-Castelvetri et al., 2020); which are widely expressed in different tissues of human body. Symptoms of COVID-19 patients include fever, cough, fatigue, headache, diarrhea, and in severe cases may lead to organ failure. Different patients have various symptoms after SARS-CoV-2 infection, and most of them develop mild to moderate illness. Current COVID-19 pandemic poses a challenge to the healthcare system, and epidemiologists have long warned that COVID-19 is very likely to become endemic or epidemic in the human population for decades to come. Therefore, tracking the human response after SARS-CoV-2 infection is critical to facilitate the medical treatment and rehabilitation of COVID-19 patients.

Previous in-depth transcriptome analysis revealed aberrant inflammatory response in COVID-19 patients(Blanco-Melo et al., 2020). We also reported inappropriate upregulation of transposable elements (TEs) especially retrotransposons upon SARS-CoV-2 infection in human cell lines and its potential harm(Yin et al., 2021). TEs are mobile DNA elements and comprise about 40% of human genome(Dewannieux et al., 2003). Four major TE classes are Long interspersed nuclear elements (LINEs), short interspersed nuclear elements (SINEs), long terminal repeats (LTRs) and DNA transposons. These are involved in many cellular processes, such as: transcriptional regulation(Percharde et al., 2018), chromatin structure organization(Fadloun et al., 2013), development and differentiation (Lu et al., 2020; Padmanabhan Nair et al., 2021). Retrotransposons are active TEs capable of “copy and paste” themselves into the human genome through RNA intermediates. Well-known retrotransposons include LINEs, SINEs and LTRs. LINEs are the most common autonomous retrotransposons, and the mobilization activity of SINEs relies on LINEs(Cordaux and Batzer, 2009; Dewannieux et al., 2003). Retrotransposons have the capacity to cause insertion, deletion and inversion in human genome and therefore their increased expression may lead to reduced genome stability(Gilbert et al., 2002; Malki et al., 2019; Newkirk et al., 2017; Symer et al., 2002). Regarding the impact of SARS-CoV-2 on human body, a recent study using peripheral blood from COVID-19 patients showed impaired transcriptional network and epigenetic profiles which might be useful for targeted treatment and provided promising hallmarks to predict clinical outcome(Bernardes et al., 2020). However, how SARS-CoV-2 impacts TEs in human body remains unclear.

Despite intensive multi-omics investigations (Bojkova et al., 2020; Delorey et al., 2021; Nie et al., 2021; Shen et al., 2020; Wu et al., 2021; Xiong et al., 2020), the impact of SARS-CoV-2 on the human body and the long-term effect of viral infection remains largely unexplored; and there is still uncertainty about continuous influence on the health of COVID-19 patients after their recovery. Notably, SARS patients were tracked after outbreak of SARS in 2003, identifying a significant incidence of sequelae, including pulmonary fibrosis and limited body function(Gomersall et al., 2004). During the current pandemic, COVID-19 patients were reported to suffer from fatigue, sleep difficulties, and anxiety/depression several months’ after recovery(Huang et al., 2021). Meanwhile, studies on COVID-19 patients showed severely impaired gut microbiota up to 3-to-6 months after recovery (Chen et al., 2021; Tian et al., 2021). Moreover, pulmonary dysfunction and ‘plasma metabolites’ remain incompletely restored 3 months after recovery (Chen et al., 2021). Understanding the progress of convalescence of COVID-19 patients is therefore valuable for clinical intervention.

Gene expression pattern and epigenetic profile of peripheral blood reflect the whole body metabolic status. In the current study, we collected peripheral blood from COVID-19 patients 3 months after recovery from COVID-19, and carried out transcriptome and DNA methylome studies (Figure 1A). Significant amounts of misregulated genes/TEs and differentially methylated regions (DMRs) were identified, indicating incomplete restoration of human body. Additionally, we identified transcriptome and epigenome signatures which will help identify the long-term impact of COVID-19 on health and provide suggestions for clinical treatment.

**Figure 1.**
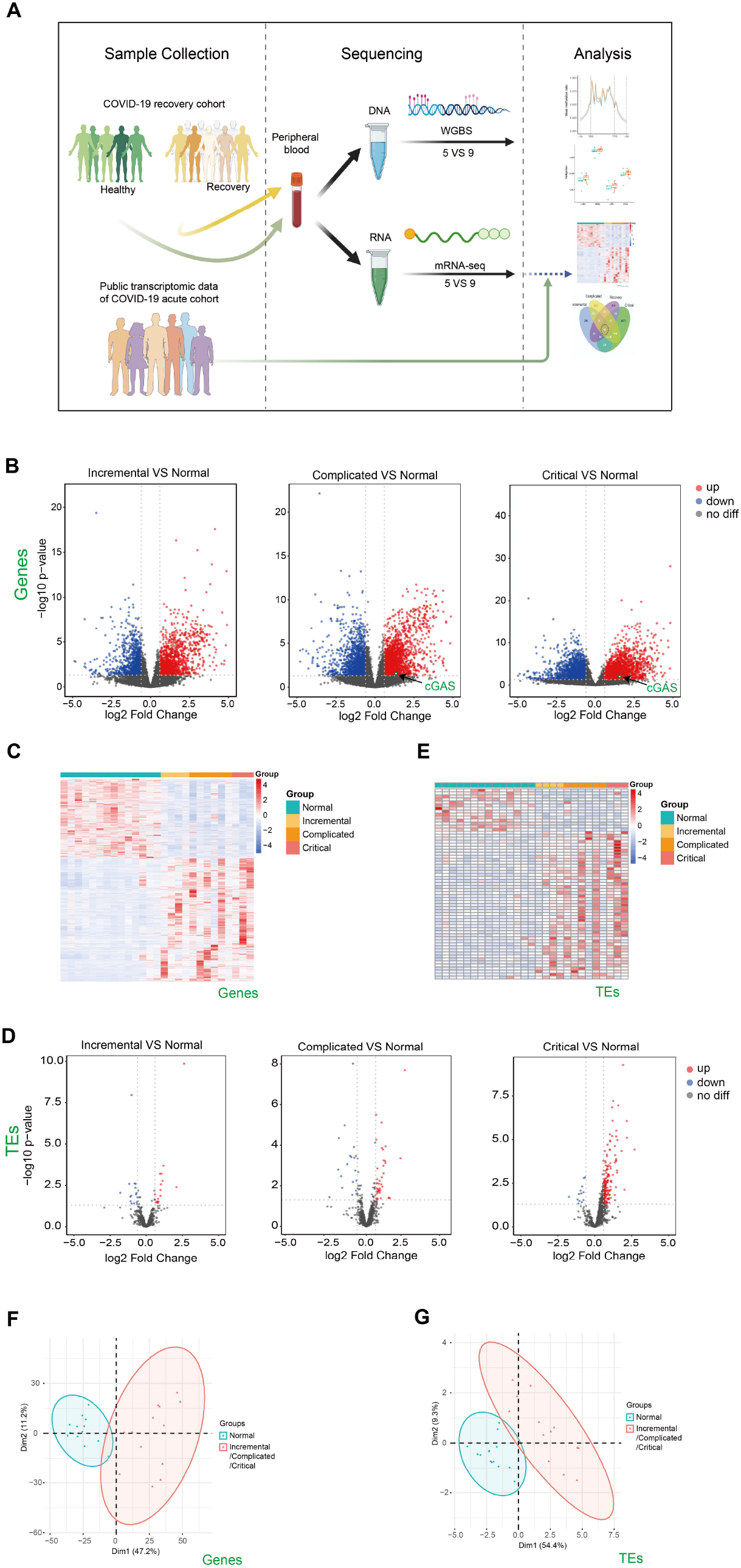
Transcriptome changes of peripheral blood samples from COVID-19 patients at acute phase of incremental, complicated, and critical stages by public RNA-seq data analysis. **(A)** Scheme illustrating our experimental design created with BioRender.com. **(B)** Volcano plots (-log_10_(p value) versus log_2_(foldchange of gene expression)) displaying transcriptome changes (|log_2_foldchange|>0.6 and p value<0.05) of COVID-19 patients at incremental, complicated, and critical stages. Upregulation of cGAS at acute phase of complicated and critical stages is indicated. **(C)** Heatmap shows DEGs at incremental, complicated, and critical stages. **(D)** Volcano plots displaying TE expression changes (|log_2_foldchange|>0.6 and p value<0.05) of COVID-19 patients at incremental, complicated, and critical stages. **(E)** Heatmap shows DETEs at incremental, complicated, and critical stages. **(F)** PCA clusters the sequenced samples by normalized counts for DEGs of incremental/complicated/critical group and control group. **(G)** PCA clusters the sequenced samples by normalized counts for DETEs of incremental/complicated/critical group and control group.

## RESULTS

### SARS-CoV-2 infection had a profound impact on transcriptome and TE activation which was positively correlated with COVID-19 severity

To identify how human body responds to SARS-CoV-2, we downloaded and analyzed public RNA-seq data of peripheral blood from COVID-19 patients at acute phase including incremental, complicated and critical stages, as well as healthy control(Bernardes et al., 2020) (Table S1). A total of 2392, 4241 and 3655 misregulated genes were identified in these three stages respectively (Figure 1B and 1C). DEGs were mainly enriched in immune response, cell cycle, and DNA repair (Figure S1A, S1B). We previously reported that expression of TEs was upregulated upon SARS-CoV-2 infection in human cell lines(Yin et al., 2021). Therefore, we examined expression of TEs in acute phase, and observed a gradual upregulation of TEs from incremental (16 upregulated) to complicated (41 upregulated) to critical (164 upregulated) stage (Figure 1D, 1E, S2), indicating that TE expression levels reflected COVID-19 severity. In agreement with identification of TE activation, we also noticed cGAS upregulation at complicated and critical stages (Figure 1B), and retrotransposon upregulation is one of the major reasons stimulating cGAS-STING pathway which may provide therapeutic targets for reducing inflammation in COVID-19 patients. PCA effectively clustered the samples by either DEGs (Figure 1F) or DETEs (Figure 1G). Collectively, our results indicated that SARS-CoV-2 infection triggered immune response and activated TEs in human body.

### Whole-blood transcriptome analysis revealed that SARS-CoV-2 impacted expression of multiple genes and alternative splicing events up to 3 months after recovery

To understand the recovery progress of human body after 3-month convalescence from SARS-CoV-2 infection, we recruited COVID-19 patients with mild/moderate symptoms and controls at similar ages in Hubei province, China. Generally, patients had a median age of 35.7 years, and control group had a median age of 32.8 years. The median follow-up time after hospital discharge was 86 days (approximately 3 months). Peripheral blood was collected for subsequent RNA-seq analysis (see Table S2 for quality control information). Generally, 425 genes were upregulated and 214 genes were downregulated (Figure 2A, 2B and 2C). PCA effectively clustered the samples by DEGs (Figure 2D). DEGs were enriched in immune response-related biological processes (Figure 2E). We then ask whether recovery group and acute phase groups have similar misregulated genes. We found that among 639 misregulated genes in recovery group, 205 genes were also misregulated in acute phase groups (Figure 2F). There were 48 genes misregulated in all four disease stages, all involved in T cell activation and other immune response-related processes (Figure 2F and S3). As expected, these 48 overlapping misregulated genes can be used to clearly discriminate COVID-19 patients from controls, no matter whether the patients were at acute or recovery stage (Figure 2G), and are therefore useful for clinical diagnosis and treatment of COVID-19.

**Figure 2.**
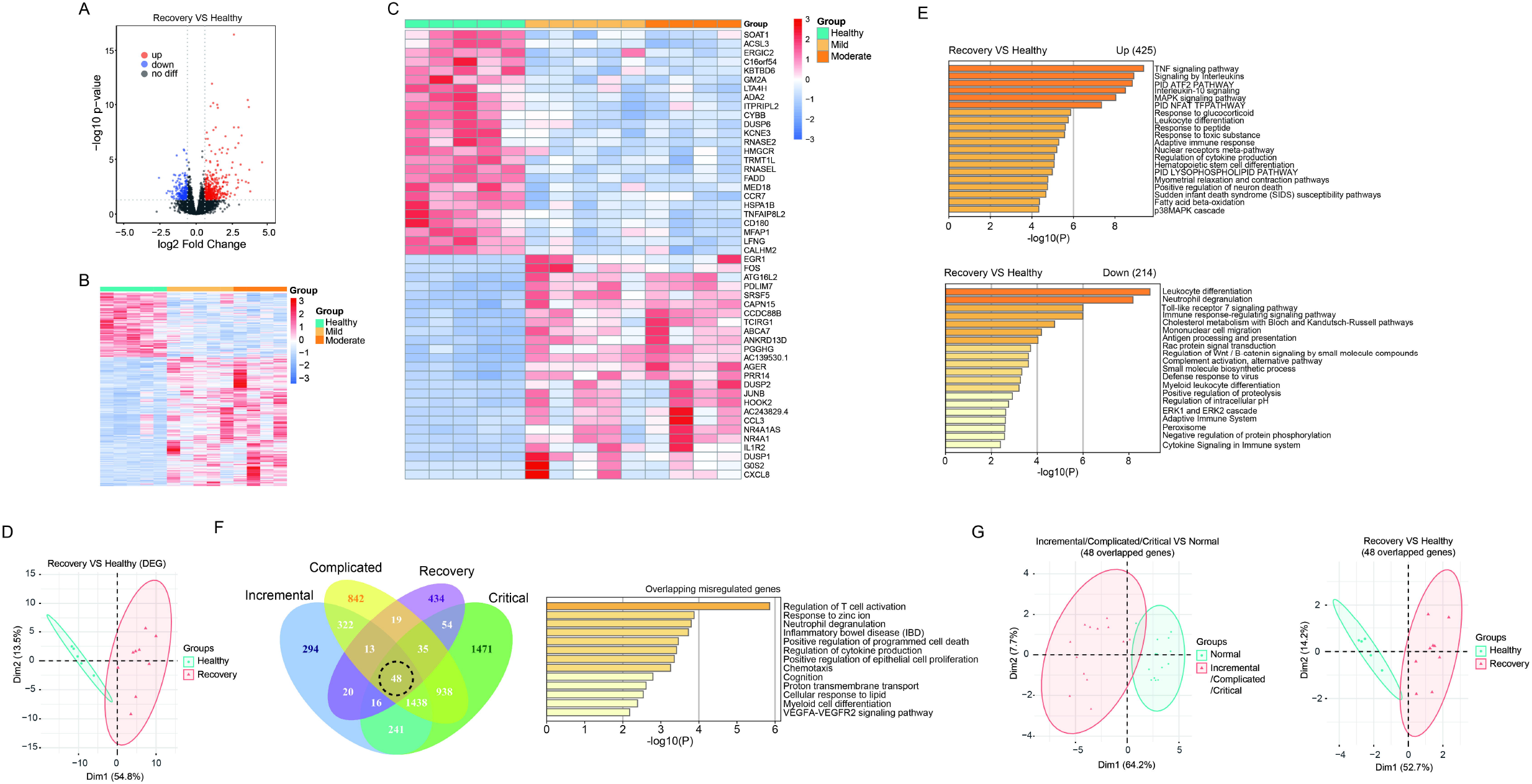
Transcriptome analysis identified gene expression changes of peripheral blood samples from COVID-19 patients after 3-month recovery. **(A)** Volcano plots (-log_10_(p value) versus log_2_(foldchange of gene expression)) displaying transcriptome changes (|log_2_foldchange|>0.6 and p value<0.05) at recovery stage. **(B)** Heatmap shows DEGs in COVID-19 patients at recovery stage. **(C)** Heatmap shows top 25 downregulated and top 25 upregulated genes at recovery stage. **(D)** PCA clusters the sequenced samples by normalized counts for DEGs of recovery group and control group. **(E)** GO analysis of upregulated/downregulated genes for functional enrichment at recovery stage by Metascape. **(E)** Venn diagram identifies 48 overlapping misregulated genes among incremental, complicated, critical stages and recovery stage. GO analysis was further performed to identify functional enrichment of the 48 genes by Metascape. **(G)** PCA of incremental/complicated/critical group and control group (left) and PCA of recovery group and control group (right), using 48 overlapped genes. PCA clusters the sequenced samples by normalized counts for 48 overlapping misregulated genes among incremental, complicated, critical stages and recovery stage.

Next, we analyzed expression changes of TEs in recovery group. When we added up all reads for each class of transposon, we observed that global expression of LINE, SINE, LTR and DNA transposon were all significantly increased (Figure 3A). There were 62 upregulated TEs with most of them belonging to SINE and LTR, while no downregulated TE subfamilies were observed (Figure 3B, 3C and Table S3). Notably, those with moderate illness seemed to have higher upregulation of DETEs than those with mild illness, indicating that TE levels reflected severity of COVID-19 at recovery stage. PCA effectively clustered the samples by DETEs (Figure 3D). Surprisingly, 56 of the 62 upregulated TEs in recovery group were absent in patients from the acute phase group, indicating that distinct TE subfamilies were activated during the recovery progress.

**Figure 3.**
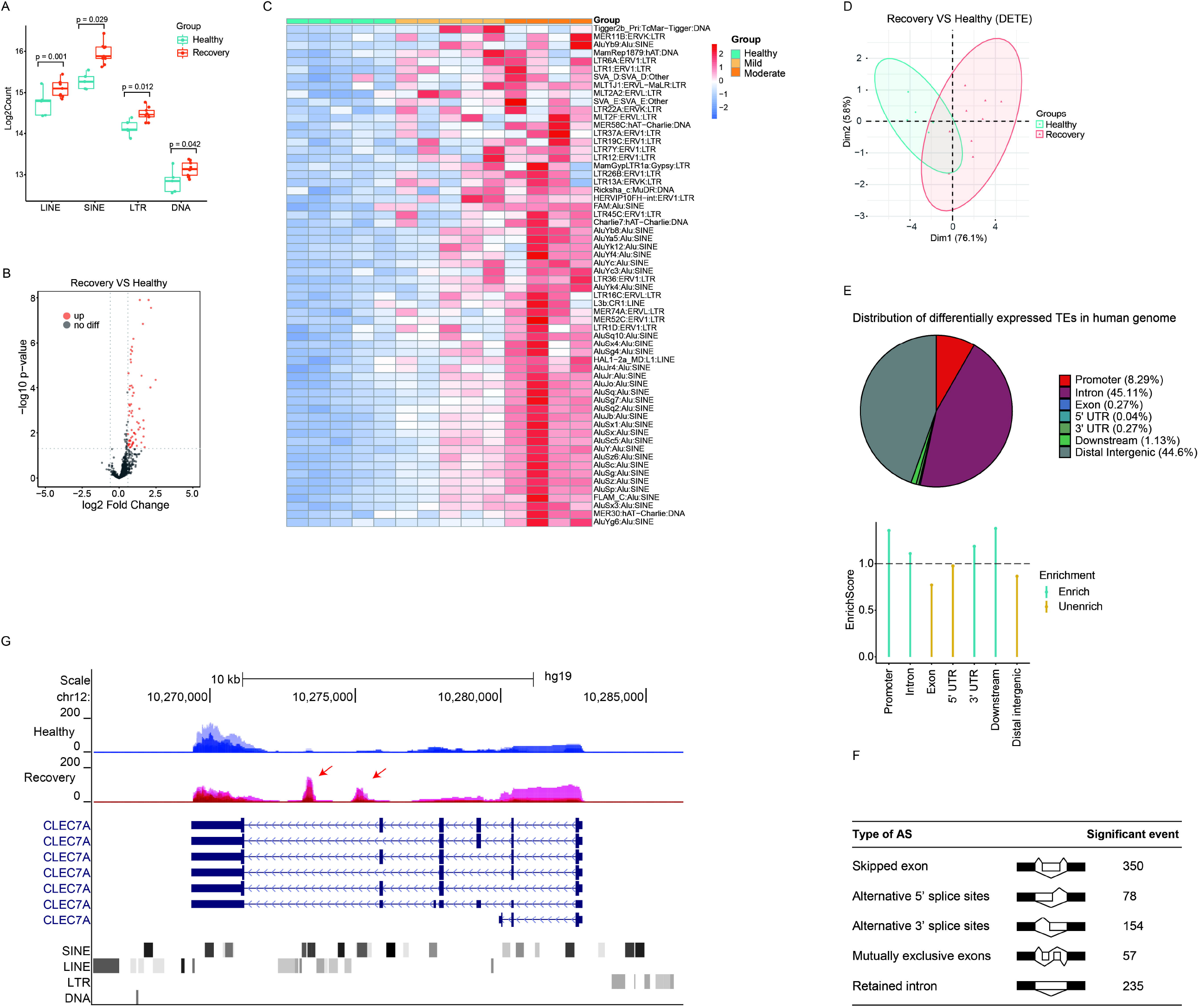
Discovery of aberrantly overexpressed TEs in peripheral blood samples from COVID-19 patients after 3-month recovery. **(A)** Boxplot displaying significantly upregulation of four major TE classes. LINE, SINE, LTR, and DNA transposons are shown, and each dot represents one sample. The median, first, and third quartiles are shown. Two-sided Wilcoxon signed-rank test were used for the comparisons. **(B)** Volcano plots (-log_10_(p value) versus log_2_(foldchange of gene expression)) displaying TE expression changes (|log_2_foldchange|>0.6 and p value<0.05) at recovery stage. **(C)** Heatmap demonstrates all 62 upregulated TEs in COVID-19 patients at recovery stage. **(D)** PCA clusters the sequenced samples by normalized counts for DETEs of recovery group and control group. **(E)** Pie chart shows the distribution of DETE in different gene features of human genome. Enrich score was calculated by (Feature ratio of DETE)/(Feature ratio of TE), chi-square tests was used for statistics, enrichment score >1 and p-value < 0.05 was defined as enrichment. **(F)** Change of alternative splicing (AS) events (FDR<0.05) was observed at recovery stage. **(G)** UCSC genome browser view of RNA-seq data demonstrates aberrantly increased TE expression from an intron of CLEC7A gene initiates a novel transcript within CLEC7A gene locus.

Different TE subfamilies were misregulated in patients from various stages may be explained by crosstalk between different TE subfamilies. Therefore, we analyzed previously reported transcriptome data of LINE1 knockdown in mouse embryonic stem cells (mESCs)(Percharde et al., 2018) and indeed found abnormally expressed subfamilies of SINE and LTR (Figure S4). Next, we analyzed the distribution of upregulated TEs in human genome and found that they were mainly enriched in promoter, intron and downstream regions (Figure 3E). Meanwhile, our transcriptome analysis identified significant changes of alternative splicing events in the recovery group (Figure 3F). Although it is unclear how TEs were activated in the recovery group, increased expression of some of these may induce generation of novel transcripts inside gene loci and impact alternative splicing patterns (Figure 3G). Besides transcriptional disturbance and enhanced TEs expression may reduce genome stability, induce inflammation, and cause age-associated disorders.

Gene expression can be regulated by DNA methylation which can be long-term memorized. Thus, we next examined the expression of DNMT and TET family genes which play key roles controlling DNA methylation status. Almost all of these showed altered expression levels in patients at recovery stage (Figure S5). This prompted us to further investigate changes of genome-wide DNA methylation in COVID-19 patients after 3 months recovery.

### Whole-blood DNA methylome analysis identified genome-wide DMRs which mainly localized at TEs’ loci between healthy and recovery groups

To study how DNA methylome was changed between healthy and recovery groups and how it may correlate with transcriptome alteration, we examined whole-blood DNA methylome by WGBS (see Table S4 for quality control information). Generally, whole-genome CG methylation levels showed no significant alterations (Figure 4A and 4B). We further analyzed promoter, exon, intron and intergenic regions and did not find significant differences between recovery group and control group (Figure 4C). Analysis of genomic regions from 2 kb upstream of transcriptional start sites (TSSs) to 2 kb downstream of transcriptional end sites (TESs) indicated no significant CG methylation changes (Figure 4D). Next, we examined loci of TEs and identified minimum alteration of global CG methylation level (Figure 4E and 4F).

**Figure 4.**
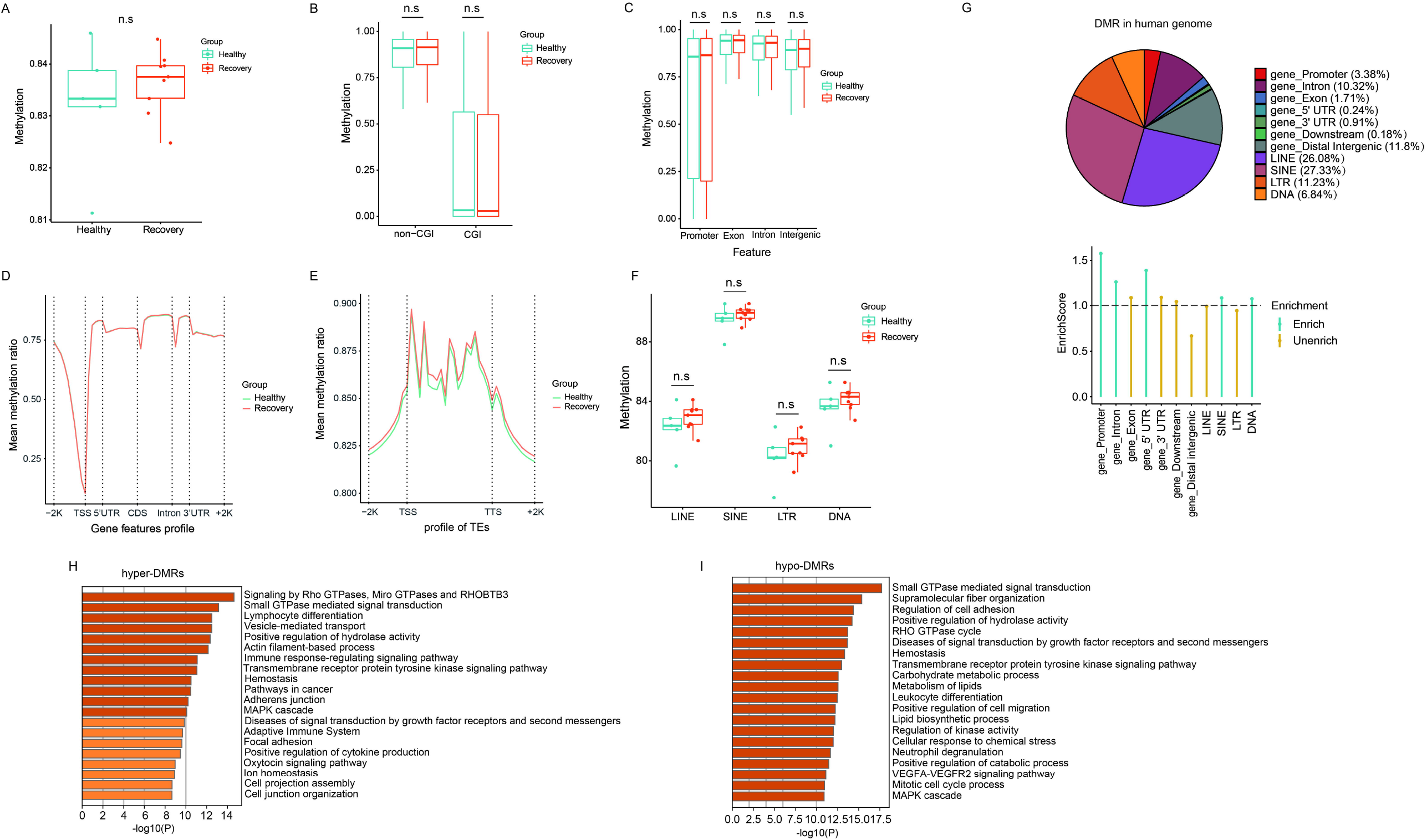
WGBS analysis of recovery and control group revealed comparable DNA methylation profiles and genome-wide DMRs which mainly localized at TE loci. **(A)** The whole mean methylation level per sample of 18M CpG sites for Healthy and Recovery group, and each dot represents one sample. Boxplot shows the mean methylation status per samples of Healthy and Recovery group, each dot represents mean methylation status of one sample. **(B)** Boxplot of mean methylation status of CGI and non-CGI of Healthy and Recovery group. **(C)** Boxplots show the mean methylation status of gene features (promoter, exon, intron and intergenic) of Healthy and Recovery group. **(D)** Methylation profile of gene features per sample. Methylation levels were measured in each 200 bp interval of a 2kb region upstream and downstream of all annotated genes. Methylation was measured in 10 equally sized bins for CDSs and introns, and 5 equally sized bins for UTRs. **(E)** Methylation profile of TEs per sample. Methylation levels were measured in each 200 bp interval of a 2kb region upstream and downstream of all annotated TEs, then for each TEs methylation levels were measured in 20 equally sized bins. **(F)** Boxplot of mean methylation status of four major TE classes. LINE, SINE, LTR, and DNA transposons are shown, and each dot represents one sample. **(G)** Pie chart demonstrates distribution of DMRs in different gene features of human genome and TE subfamilies. Enrich score was calculated by (Feature ratio of DMR)/(Feature ratio of reference genome), sliding windows was made across reference genome with 500 bps consistent with our call DMR strategy. Chi-square tests was used for statistics, enrichment score >1 and p-value < 0.05 was defined as enrichment. **(H)** Functional enrichment of genes with hyper-DMRs identified at promoter/gene body by Metascape. **(I)** Functional enrichment of genes with hypo-DMRs identified at promoter/gene body by Metascape. For above boxplots, the median, first, and third quartiles are shown. Two-sided Wilcoxon signed-rank test were used for all the comparisons.

To explore changes of CG methylation pattern between healthy and recovery groups, we analyzed DMRs and identified 18516 DMRs in total (absolute methylation mean difference > 10% and q-value < 0.05). 8724 DMRs had hypo-methylation (hypo-DMRs) and 9792 DMRs had hyper-methylation (hyper-DMRs) in the recovery group. Identified DMRs were mainly enriched at gene promoter, intron, and certain TE regions (Figure 4G). GO analysis showed that hyper-DMRs and hypo-DMRs in gene promoter/body regions were involved in signal pathways, immune response, and metabolism (Figure 4H and 4I).

CG methylation at different genomic loci may play different regulatory roles. Without annotation of TE loci, the distribution of identified DMRs in human genome were as follows: 9.91% in promoters, 40.64% in gene bodies, 49.45% in intergenic and other regions (Figure 5A). We then examined expression levels of identified genes with DMRs at promoters or gene bodies. We found that 9 hyper-DMRs at promoters were associated with downregulated genes, while 28 hypo-DMRs at promoters were associated with upregulated genes (Figure 5B, Table S5), and these genes were mainly involved in immune responses (Figure 5C). Meanwhile, 27 hypo-DMRs at gene bodies were associated with gene downregulation, 13 hyper-DMRs at gene bodies were associated with gene upregulation (Figure 5D), mainly involved in stress response and related signaling pathways (Figure 5E).

**Figure 5.**
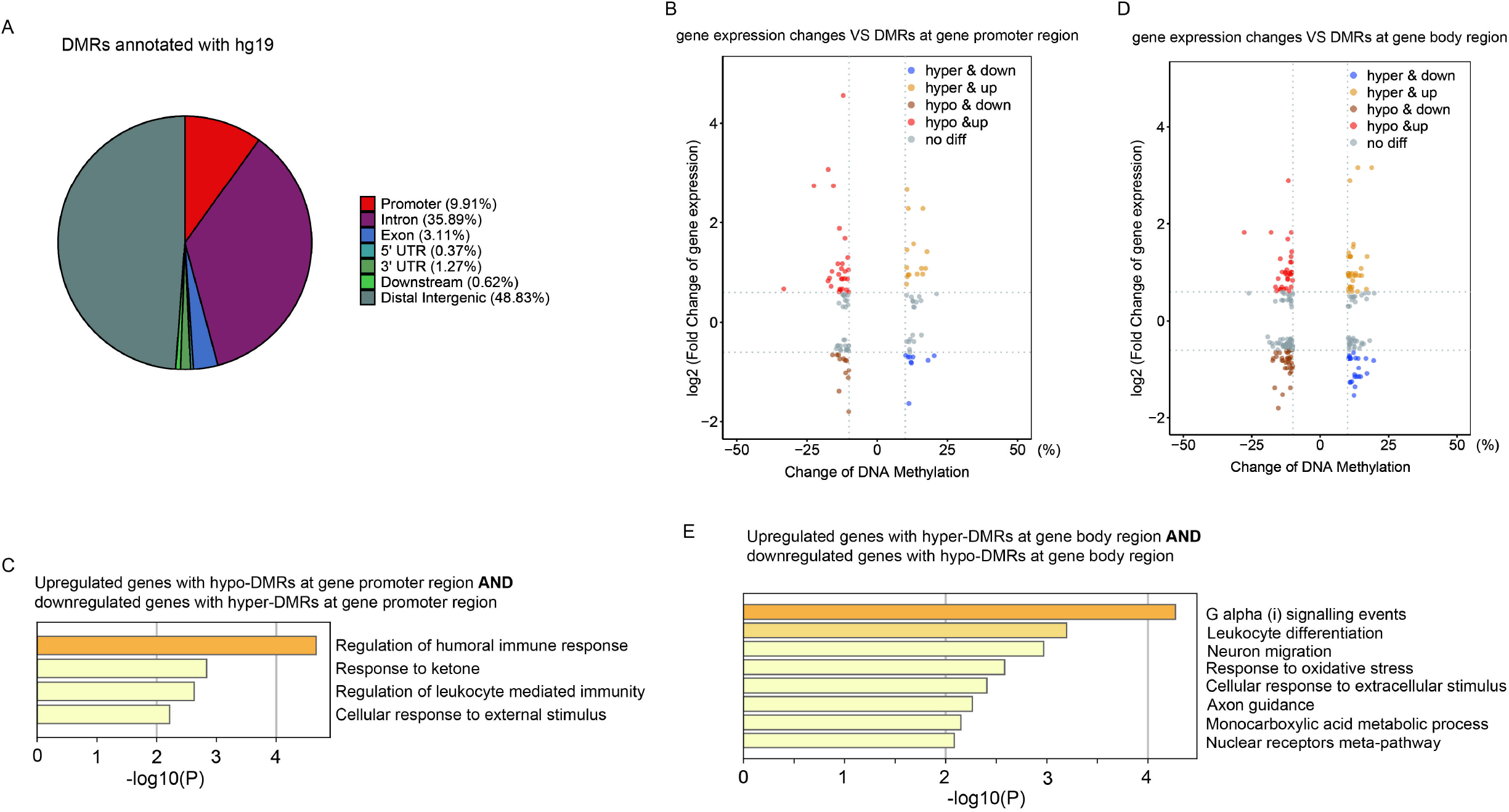
Aberrant DNA methylation at gene loci may be involved in immune response-related gene misregulation. **(A)** Pie chart (without annotation of TEs) demonstrates distribution of DMRs in human genome. **(B)** Scatter plot of the relationship between DNA methylation differences at gene promoter and their expression differences. **(C)** Functional enrichment of upregulated/downregulated genes with hypo/hyper-DMRs in gene promoter regions by Metascape. **(D)** Scatter plot of the relationship between DNA methylation differences at gene body and their expression differences. **(E)** Functional enrichment of upregulated/downregulated genes with hypo/hyper-DMRs in gene body regions by Metascape.

Next, we focused on DMRs between healthy and recovery groups within TE loci, and identified 13233 DMRs. The percentages of these DMRs were as follows: 36.48% in LINE, 38.23% in SINE, 15.71% in LTR, and 9.58% in DNA transposon (Figure 6A). Subfamilies of SINE with altered DNA methylation were mainly Alu; subfamilies of LINE were mainly LINE1; subfamilies of LTR were mainly ERVL-MalR; and subfamilies of DNA transposon were mainly hAT-Charlie (Figure 6B). We then analyzed expression changes of different TEs with altered DNA methylation and identified upregulated expression of TEs (which mainly localized at introns) associated with both hyper-and hypo-DMRs in TEs (Figure 6C). Upregulation of certain TEs without hypo-DMRs could be explained by aberrant expression of specific transcriptional regulators. DMRs at TE loci mainly distributed at intergenic regions, intron and promoter regions (Figure 6D). To explore the potential impact of DNA methylation changes of intergenic TEs on gene expression, we identified adjacent genes of intergenic TEs with altered DNA methylation and calculated total normalized gene counts for each sample. Interestingly, we observed significant upregulation of genes adjacent to TEs with increased DNA methylation, suggesting that these intergenic TEs act as distal gene silencers (Figure 6E). Further analysis revealed that 19 hypo-DMRs annotated at TE loci were located at promoter of upregulated genes, and 9 hyper-DMRs annotated at TE loci were located at promoter of downregulated genes (Figure 6F). GO analysis showed their function in ERK signaling regulation and T cell activation (Figure 6G). Furthermore, we identified DMRs at promoter regions were annotated as various TE subfamilies, although most of those TE subfamilies were not more enriched in DMRs at gene promoter region relative to other regions in human genome (Figure 6H). One example is SIK1 gene which was upregulated in recovery group and its promoter contains hypo-DMR overlapped with SINE (Figure 6I). These results indicated that TEs with altered DNA methylation may function as regulatory elements for adjacent gene expression.

**Figure 6.**
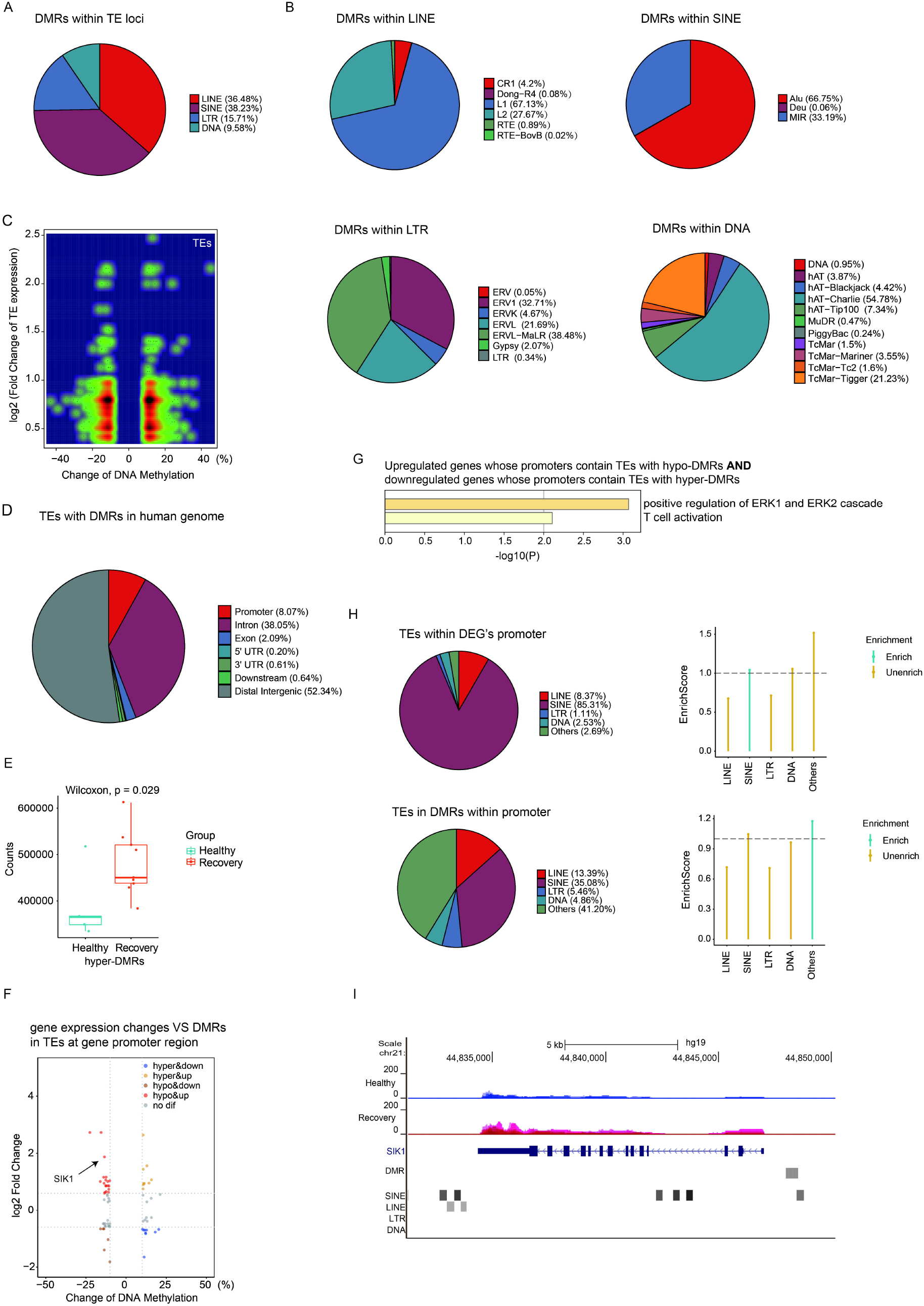
Aberrant DNA methylation at TE loci may be involved in gene regulation. **(A)** Pie chart demonstrates distribution of DMRs in TE subfamilies. **(B)** Pie chart demonstrates distribution of DMRs in LINE/SINE/LTR/DNA transposons. **(C)** Density scatter plot demonstrates relationship between DNA methylation differences of TEs and their expression differences. **(D)** Pie chart (without annotation of TEs) demonstrates distribution of TEs with DMRs in human genome. **(E)** Calculation of total gene counts adjacent to TEs with hyper-DMRs in healthy and recovery group. The median, first, and third quartiles are shown in boxplots. Two-sided Wilcoxon signed-rank test were used for all the comparisons. **(F)** Scatter plot of the relationship between DNA methylation differences at TEs and expression differences of genes with promoters containing TEs with DMRs. **(G)** Functional enrichment of upregulated/downregulated genes whose promoters contain TEs with hypo/hyper-DMRs by Metascape. **(H)** The upper pie chart demonstrates the distribution of TEs overlapped with DEG’s promoters in TE subfamilies. Enrich score was calculated by (TE subfamily ratio of TEs overlapped with DEG’s promoters)/(TE subfamily ratio of TEs overlapped with all gene’s promoters). The bottom pie chart shows the distribution of DMRs overlapped with gene’s promoters in TE subfamilies. Enrich score was calculated by (TE subfamily ratio of DMRs overlapped with gene’s promoters)/(TE subfamily ratio of gene promoters). Sliding windows was made across gene promoters with 500 bps consistent with our call DMR strategy. Chi-square tests was used for statistics, enrichment score >1 and p-value < 0.05 was defined as enrichment. **(I)** UCSC genome browser view of RNA-seq data demonstrated increased SIK1 expression in recovery group and position of hypo-DMR in SIK1 promoter.

## DISCUSSION

Our previous study on SARS-CoV-2 infected human cell lines showed viral infection-induced gene misregulation and upregulation of TEs (Yin et al., 2021). However, how TEs behave in human body remain elusive. To identify how transcriptional program responds to SARS-CoV-2 in the human body, we downloaded and analyzed a public RNA-seq dataset of peripheral blood samples from COVID-19 patients at acute phase(Bernardes et al., 2020). As anticipated, we observed both misregulation of genes and aberrantly activation of TEs.

Despite extensive investigations, the outcome of SARS-CoV-2 infection, long-term health consequences (Nabavi, 2020) and long-term recovery progress of COVID-19 patients remains elusive. Majority of the COVID-19-related DMRs are near the gene promoter regions and were hypo-methylated even though the global methylation level remains similar between healthy control and COVID-19 patients (Balnis et al., 2021). However, currently reported results on DNA methylome of COVID-19 patients (Balnis et al., 2021; Bernardes et al., 2020; Li et al., 2021) all depended on 450K or 850K methylation array which covered only a small percent of CpG sites (1% to 3 %) and couldn’t be used to obtain DNA methylation information of transposable elements. Our WGBS data produced nearly whole-genome CpG sites coverage (coverages of all samples are around 80%) which is valuable for genome-wide identification of differentially methylated regions, especially for transposable elements. Here, we asked whether 3 months was enough to restore the transcriptome and DNA methylation of the patient’s peripheral blood cells to normal. Based on our results, even though the global methylation level remains the same between healthy control and 3-month recovered patients, the overall transcriptome or epigenome profile of peripheral blood samples from the recovery group is still sufficiently different from the control group. Notably, subfamilies of TEs remained upregulated 3 months after recovery, suggesting that a longer timeframe may be needed to return to normal levels. Retrotransposon can encode proteins and form retrovirus-like particles (Grow et al., 2015), so it is possible that some virus-like particles visualized by electron microscopy (EM) in COVID-19 patients (Yao et al., 2021) may derive from TEs like LTRs due to their enhanced expression rather than SARS-CoV-2. While some aberrant gene expression can be interpreted by DNA methylation changes, other mechanisms undermining the transcriptional network in human body need further exploration. Our WGBS analysis showed no significant changes of global DNA methylation and no significant changes of DNA methylation at TE subtypes, and this may be caused by bulk and heterogeneous levels of cells. Interestingly, we still found 18516 DMRs between healthy and recovery group, and 13233 DMRs were within TE loci. This supported altered TE expression and changes of DNA methylation at TE loci. It should be noted that there was contribution of cell population differences between healthy and recovery groups in identified genes/TEs which showed differential expression patterns or DNA methylation patterns. For example, healthy and recovery individuals may have different circulating memory T cells/exhausted T cell features. However, at least part of those changes at TE loci should be due to TE activation in acute and recovered patients. One supporting evidence of TE activation is that our previous study on human cell lines showed upregulation of TE upon SARS-CoV-2 infection (Yin et al., 2021). Another important supporting evidence is that our analysis on RNA-seq data of acute patients showed activation of cGAS-STING pathway (see Figure 1B for cGAS upregulation in acute patients) which can be triggered by upregulation of retrotransposon-derived cytoplasmic DNA (Decout et al., 2021). We did not identify significant activation of cGAS gene in recovered patients, probably because TE activation is not severe enough to activate cGAS-STING pathway. Besides transcriptomes and DNA methylomes, whether COVID-19 leaves other irreversible sequelae requires further investigation, such as telomere length (TL) and mitochondrial DNA (mtDNA) copy number, which are associated with many diseases including cardiovascular diseases, psychiatric disorder, cancers and inflammatory diseases (Shay, 2016; Sun and St John, 2016).

Our study reveals genes with aberrant expression and genomic regions with altered epigenetic modification in COVID-19 convalescent patients 3 months after recovery. Our results support long-term disease stage marked by overactivated immune response. Besides, this report provides potential genomic targets to facilitate convalescence of COVID-19 patients. Moreover, we provide potential transcriptional and epigenetic signatures to track SARS-CoV-2 infection history, identify profound viral impact on human cells and reveal long COVID-19 risks. However, due to genetic mutations of SARS-CoV-2 variants, gender, health, age of patients and other factors, transcriptional and epigenetic changes may vary and need further validation and investigations.

In summary, we examined the transcriptomes and DNA methylomes of COVID-19 patients 3 months after recovery, and noticed that both genes and TEs were impacted at transcription and DNA methylation level (Figure 7). Misregulated genes were involved in immune response and other biological functions; while TEs in intron and other regions were specifically activated which may disrupt transcription process and genome integrity and induce inflammation. Furthermore, DNA methylome analysis showed that genes with DMRs were also involved in immune response-related processes; and differentially methylated promoter and distal intergenic region may play important roles in gene regulation. Finally, altered CG methylation can indirectly impact gene expression and may play regulatory roles in stress, illness, and ageing. Further studies are needed to track changes of transcriptome and DNA methylome of COVID-19 patients for longer time to identify how long is required for full recovery.

**Figure 7.**
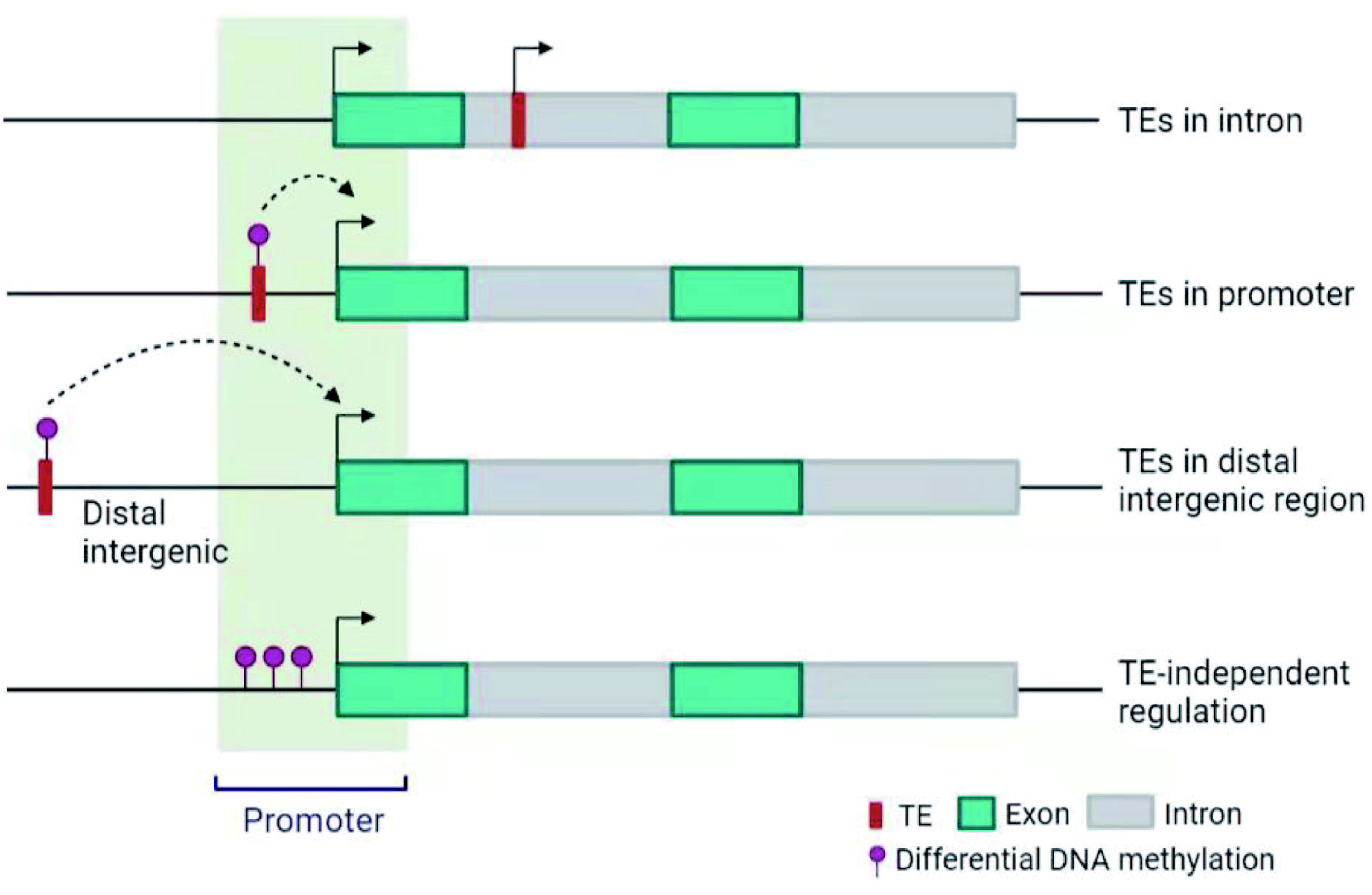
Scheme illustrating differential gene expression and DNA methylation in COVID-19 recovered patients created with BioRender.com. Gene expression is regulated in TE-dependent and -independent pathway. TEs in promoter and distal intergenic region may regulate gene expression through DNA methylation.

## MATERIALS AND METHODS

### Patient recruitment and blood sample collection

Participants were recruited from Huazhong University of Science and Technology. Inclusion criteria for COVID-19 patients included age from 25 to 45 years, having detailed medical records of hospitalization and discharge. Diagnosis of COVID-19 was determined by the New Coronavirus Pneumonia Prevention and Control Program (7th edition) published by the National Health Commission of China(http://www.nhc.gov.cn/xcs/zhengcwj/202003/46c9294a7dfe4cef80dc7f5912eb1989.shtml). Exclusion criteria included asymptomatic cases, taking antibiotic within two months, gastrointestinal diseases, and severe basic diseases. Healthy controls were recruited during regular physical check-ups in the same hospital with none of them received antibiotics within two months before collection of blood samples. Ethical approval for the study was obtained by the Ethics

Committee for Clinical Research of Reproductive Medicine Center, Tongji Medical College, Huazhong University of Science and Technology. All participants included in the study gave informed consent. 9 male patients at recovery stage and 5 male healthy controls were recruited for this study. Blood (5mL) was withdrawn from each patient into an Ethylenediaminetetraacetic acid (EDTA)-K2 tube to decelerate blood coagulation. Total RNA was isolated from 2.5ml blood using total RNA isolation kit from SIMGEN. Isolated total RNA and remaining blood sample were subjected to mRNA sequencing and whole-genome bisulfite sequencing (WGBS) by Annoroad Gene Technology Co. Ltd (Beijing).

### mRNA isolation for sequencing

Total RNA was used as input material for the RNA sample preparations. Sequencing libraries were generated using NEBNext Ultra RNA Library Prep Kit for Illumina (#E7530L, NEB, USA) following the manufacturer’s recommendations and index codes were added to attribute sequences to each sample. The clustering of the index-coded samples was performed on a cBot cluster generation system using HiSeq PE Cluster Kit v4-cBot-HS (Illumina) according to the manufacturer’s instructions. After cluster generation, the libraries were sequenced on an Illumina HiSeq 2500 system. Approximately 25M of paired-end reads (150bp ×2) for each sample are generated.

### DNA isolation for methylation profiling

For constructing WGBS libraries, the genomic DNA was fragmented to a mean size of 350 bp, followed by blunt-ending, dA-Tailing, and adaptor ligation. Insert fragments with different sizes were excised from a 2% agarose gel and purified using the QIAquick Gel Extraction Kit (QIAGEN). Purified DNA was bisulfite converted using the EZ DNA Methylation-Gold Kit (#D5006, ZYMO Research, CA, USA) and PCR amplified. The WGBS libraries were sequenced at 20×depth on an Illumina HiSeq 2500 system as paired-end reads(150bp ×2). Approximately 15 to 20 × mean coverage was generated for each sample.

### RNA-seq data processing

Raw reads were processed with Trim Galore (v0.6.4) to remove adaptor sequences and poor quality bases with ‘--q 20 --phred33 --stringency 5 --length 20 –paired’. To include as many non-uniquely mapped reads as possible, trimmed reads were firstly aligned to human genome (hg19) by STAR (v2.7.5b) (Dobin et al., 2013) with default settings including parameters ‘-- winAnchorMultimapmax 2000 --outFilterMultimapNmax 1000’. SAMtools (v1.3.1) was used to sort bam files by genomic coordination and make a bam file index. RSEM (v1.2.28) (Li and Dewey, 2011) was used to calculate FPKM value of genes. TEtranscript(Jin and Hammell, 2018) with default parameters was used to get counts for different transposable elements. UCSC genome browser was used for snapshots of transcriptome. R package Deseq2 (v1.28.1) (Love et al., 2014) was used to obtain differentially expressed genes (DEGs) and differentially expressed TEs (DETEs). Principal component analysis (PCA) was performed using DESeq2 normalized counts for DEGs/DETEs. Metascape (Zhou et al., 2019) was used to visualize functional profiles of genes and gene clusters. rMATS (v4.1.1) (Shen et al., 2014) was used to identify alternative splicing events with “--readLength 150” and other default parameters. Graphs were created by R. Images were organized by Adobe Illustrator.

### WGBS data processing and quality control

Raw reads were processed with Trim Galore (v0.6.4) to remove adaptor sequences and poor quality bases with ‘--q 20 --phred33 --stringency 5 --length 20 --paired’. Trimmed reads were then aligned to human genome (hg19) by Bismark (v0.22.3) (Krueger and Andrews, 2011) using the parameters “-p 6 --parallel 1 -N 0 -L 20 --quiet --un --ambiguous --bam”. SAMtools (v1.3.1) was used to sort bam files by genomic coordination and make a bam file index. PCR duplicates were removed using Picard (v2.23.3). The methylation ratio at each CpG site was constructed using bismark_methylation_extractor model with the parameters “-p --comprehensive --no_overlap -- bedgraph –counts --report --cytosine_report --gzip –buffer --size 30G”. For all samples, the average bisulfite conversion success ratio is >99.2%, alignment ratio is around 80% of read pairs aligning uniquely, and duplication rate is <5%. For each CpG sites, methylation levels were calculated by (methylation reads/total coverage reads). For more robust analysis, we applied the minimum threshold 3× coverage and also selected CpGs that all samples had their methylation levels. This screening process gave 18 M of CpGs with confident methylation levels. Methylation profiles were calculated by deeptools (v3.5.1) (Ramirez et al., 2014).

### Differentially methylated regions (DMRs) by methylKit

The R package methylKit (v1.14.2) (Akalin et al., 2012) was used to identify DMRs between healthy and recovery groups. The methylation levels at CpG sites were firstly calculated by “methRead” function with mincov=3. Methylation across the genome was tiled with the ‘tileMethylCounts’ function using the parameters “win.size=500, step.size=500, cov.bases =5”, then ‘unite’ function was used to unite tiled regions with the “destrand=TRUE” parameter. At last, “calculateDiffMeth” function was used to calculate DMRs. DMRs with a minimum of 3 CpG sites and absolute methylation mean difference > 10% and q-value < 0.05 were used for further analysis. DMRs were annotated by R package “ChIPseeker” (v1.24.0).

### Statistical methods

Plotting and statistical tests were performed using R (v4.0.2). All statistical tests performed in this study were two-sided. Box plots were generated using the R packages “ggplot2” (v3.3.2) and “ggpubr” (v0.4.0) to show median, first and third quartiles, and outliers were shown if outside the 1.5× interquartile range. A two-sided Wilcoxon signed-rank test was used to assess differences between two groups. Enrichment scores were analyzed using chi-square tests, enrichment score >1 and p-value < 0.05 was defined as enrichment.

## Data Availability

Public RNA-seq data of peripheral blood samples from COVID-19 patients at incremental, complicated, and critical stages were downloaded from GEO database (GSE161777). Public RNA-seq data of mESCs with LINE1 knockdown were downloaded from GEO database (GSE100939). The bulk RNA-seq and WGBS data generated during this study is available in Genome Sequence Archive (GSA) for human data repository of National Genomics Data Center (BioProject No. PRJCA006301, https://ngdc.cncb.ac.cn/bioproject/browse/PRJCA006301).

## Author contributions

Li-quan Zhou, Ximiao He, Hong-gang Li and Cheng-liang Xiong conceived and designed the project. Ying Yin, Qing Tian, Zhen Ye and Tian-qing Meng collected the blood samples. Ying Yin and Xiao-zhao Liu analyzed the data and wrote the manuscript. Yi-xian Fan and Gong-hong Wei helped with data analysis. Li-quan Zhou, Ximiao He and Hong-gang Li revised the manuscript. All authors have read and approved the final manuscript.

## Abbreviations

WGBS: whole-genome bisulfite sequencing
DMR: differentially methylated region
TEs: transposable elements
LINEs: long interspersed nuclear elements
SINEs: short interspersed nuclear elements
LTRs: long terminal repeats
DNAs: DNA transposons
DEGs: differentially expressed genes
DETEs: differentially expressed TEs

## Compliance and ethics

The authors declare no competing interests. The studies involving human participants were reviewed and approved by Ethics Committee for Clinical Research of Reproductive Medicine Center, Tongji Medical College, Huazhong University of Science and Technology. The patients/participants provided their written informed consent to participate in this study.

## Acknowledgements

The authors would like to express their thanks to Drs. Hui Zhou, Xing-jie Zhang, and Zhen-Yu Zhong for their assistance on this study. This work was supported by the National Key R&D Program of China [2018YFC1004500 and 2018YFC1004000], HUST COVID-19 Rapid Response Call [2020kfyXGYJ057], the National Natural Science Foundation of China [NSFC 31771661, 32000488], Hubei Province Natural Science Foundation [2020CFB817] and program for HUST Academic Frontier Youth Team.

