## Supplementary materials for "Transcriptome and DNA methylome analysis of peripheral blood samples reveals incomplete restoration and transposable element activation after 3-month recovery of COVID-19"

Table S1. Clinical Characteristics of COVID-19 Patients at Incremental, Complicated and Critical stages of acute phase from GSE161777 (previously published).

| No. | Gender | Ethnicity | Age (yrs) | Disease Severity | Sampling day since symptom onset |
| --- | --- | --- | --- | --- | --- |
| Incremental 1 | Male | caucasian | 55-59 | Incremental | 19 |
| Incremental 2 | Male | caucasian | 60-64 | Incremental | 9 |
| Incremental 3 | Male | caucasian | 45-49 | Incremental | 6 |
| Incremental 4 | Male | caucasian | 70-74 | Incremental | 14 |
| Complicated 1 | Male | caucasian | 60-64 | Complicated | 11 |
| Complicated 2 | Male | caucasian | 45-49 | Complicated | 8 |
| Complicated 3 | Male | caucasian | 60-64 | Complicated | 18 |
| Complicated 4 | Male | caucasian | 70-74 | Complicated | 9 |
| Complicated 5 | Male | caucasian | 70-74 | Complicated | 16 |
| Complicated 6 | Female | caucasian | 50-54 | Complicated | 36 |
| Critical 1 | Male | caucasian | 55-59 | Critical | 21 |
| Critical 2 | Male | caucasian | 60-64 | Critical | 12 |
| Critical 3 | Female | caucasian | 50-54 | Critical | 29 |

Table S2. RNA-seq quality control information of Healthy and Recovery Groups.

| Sample | Average read length | Total reads | Mapped reads (%) |
| --- | --- | --- | --- |
| H1 | 297 | 22993855 | 97.99 |
| H2 | 297 | 24665534 | 97.52 |
| H3 | 298 | 24091657 | 97.09 |
| H4 | 297 | 23706405 | 96.68 |
| H5 | 298 | 23440276 | 96.60 |
| R1 | 295 | 25638270 | 97.98 |
| R2 | 297 | 23833486 | 97.61 |
| R3 | 296 | 24684989 | 97.88 |
| R4 | 298 | 23155208 | 96.56 |
| R5 | 298 | 22523213 | 96.91 |
| R6 | 298 | 23114785 | 96.98 |
| R7 | 298 | 22875701 | 97.14 |
| R8 | 298 | 22589723 | 97.11 |
| R9 | 297 | 23621946 | 96.80 |

Table S3. Normalized counts and log_2_FoldChange of expression in differentially expressed TEs between Healthy control and Recovery patients.

Please see attached Excel file.

Table S4. WGBS quality control information of Healthy and Recovery Groups.

| Sample | Mapping Rate (%) | Mean Mapping Quality | Mean Coverage |
| --- | --- | --- | --- |
| H1 | 80.30 | 35.45 | 14.54 |
| H2 | 79.80 | 35.44 | 14.29 |
| H3 | 81.00 | 35.55 | 17.53 |
| H4 | 82.00 | 35.59 | 14.48 |
| H5 | 81.30 | 34.91 | 23.87 |
| R1 | 81.40 | 35.46 | 17.89 |
| R2 | 80.20 | 35.34 | 14.47 |
| R3 | 80.80 | 35.31 | 15.51 |
| R4 | 81.70 | 35.54 | 14.19 |
| R5 | 80.70 | 35.56 | 13.84 |
| R6 | 80.20 | 35.40 | 13.91 |
| R7 | 81.00 | 35.52 | 13.92 |
| R8 | 81.40 | 35.63 | 14.63 |
| R9 | 79.80 | 35.43 | 28.47 |

Table S5. List of 9 hyper-DMRs at promoters associated with down-regulated gene and 28 hypo-DMRs at promoters associated with up-regulated gene.

| Hyper-DMR and  downregulated | Hypo-DMR and upregulated | |
| --- | --- | --- |
| LFNG | GDPD5 | TSPOAP1 |
| CCR7 | GDPD5 | LUC7L |
| OSBPL10 | RBPMS2 | PABPC1L |
| SCIMP | KY | ARRDC2 |
| NUAK2 | MOB2 | MAP3K8 |
| NUAK2 | TANGO2 | DNAJC4 |
| NUAK2 | RHD | PILRB |
| CD163 | CR1L | MXD4 |
| ZNF468 | CFAP45 | IL1B |
|  | CFAP45 | FOSB |
|  | NUTM2A-AS1 | GAMT |
|  | ST6 | WBP2 |
|  | CCL4 | NEIL1 |
|  | LOC728392 | SIK1 |

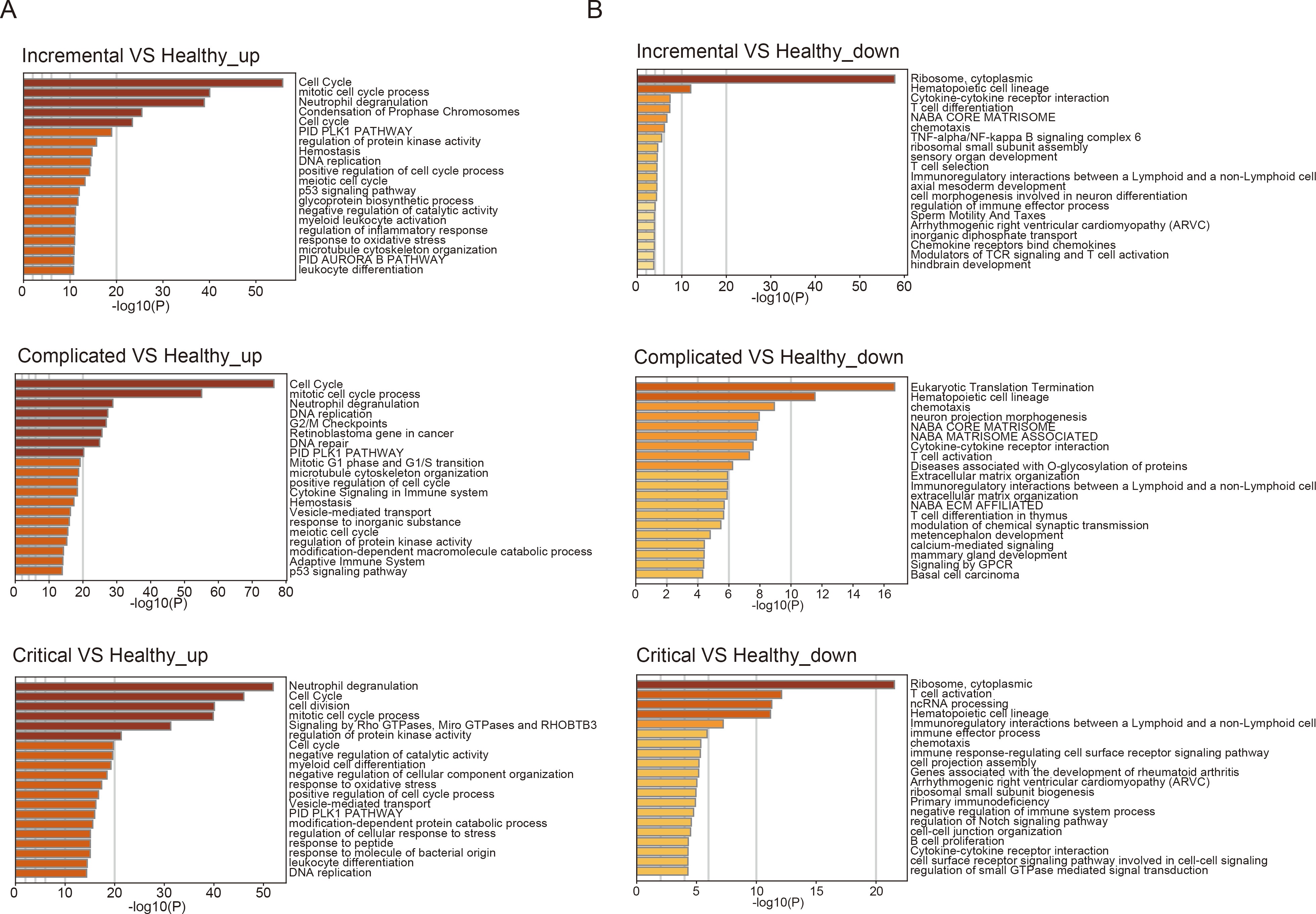

**Fig. S1.** GO analysis of upregulated(A) and downregulated genes (B) in incremental, complicated and critical stages of acute phase by Metascape.

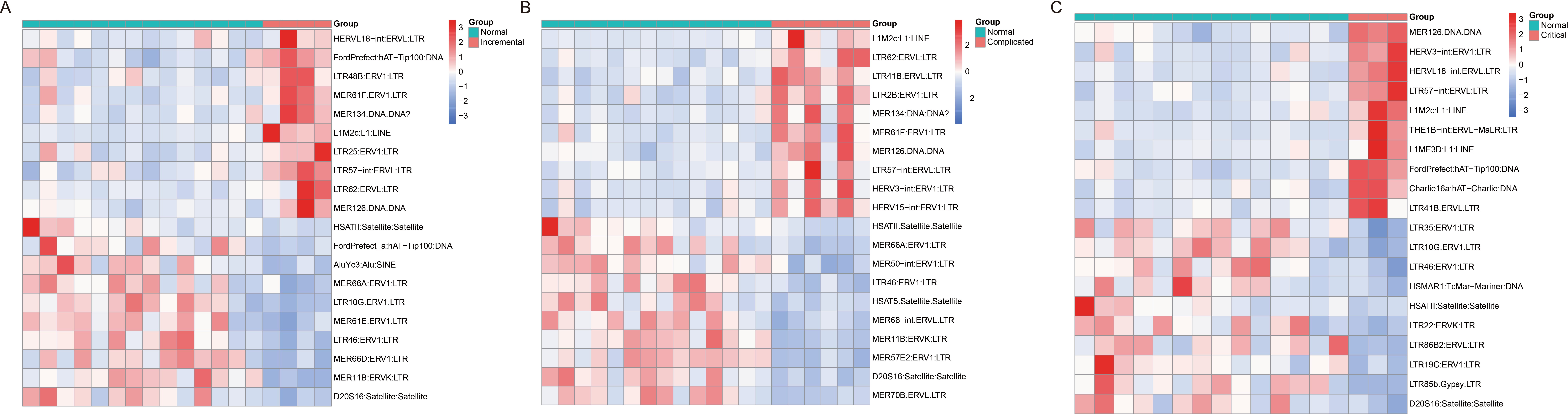

**Fig. S2.** Heatmap displaying top 10 upregulated and top 10 downregulated TEs in COVID-19 patients at incremental (A), complicated (B) and critical (C) stages of acute phase.

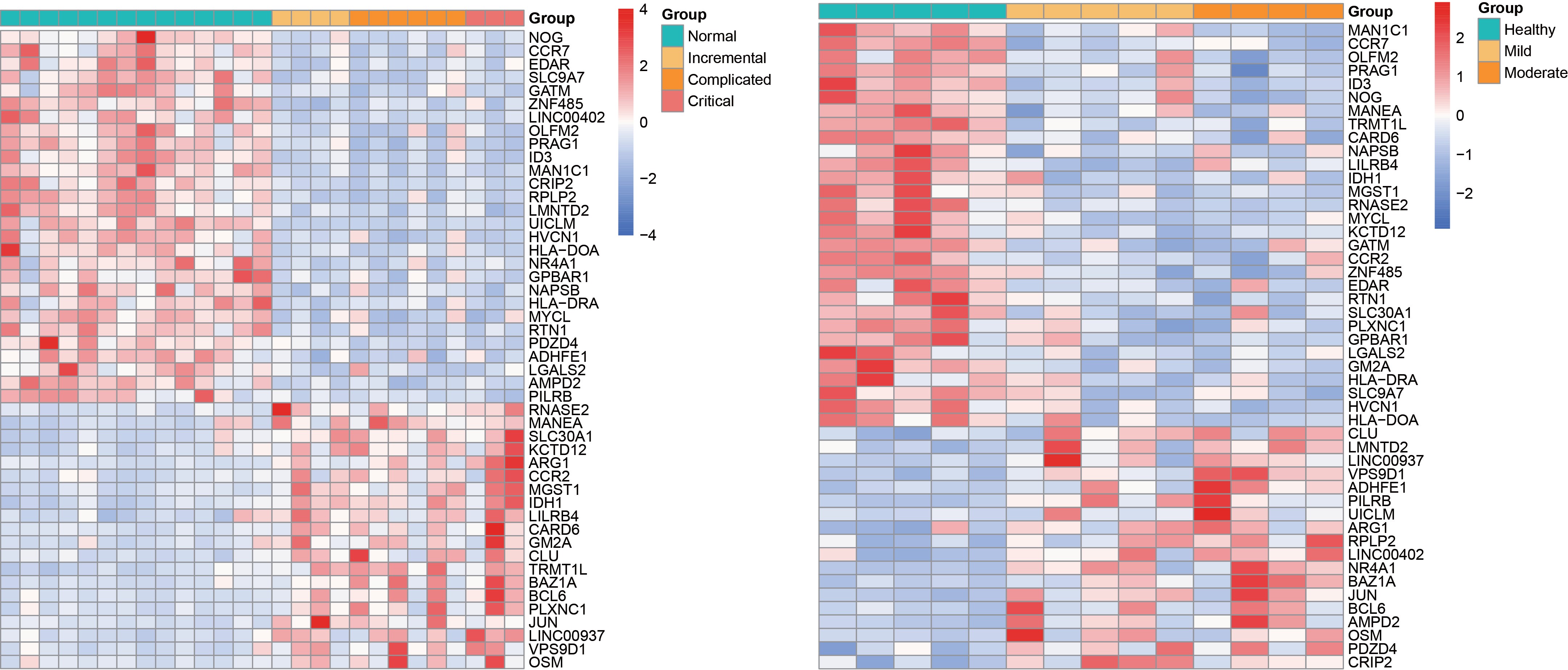

**Fig. S3.** Heatmap displaying 48 overlapping misregulated genes among acute phase (incremental, complicated, critical stages) and recovery stage (Mild and Moderate illness).

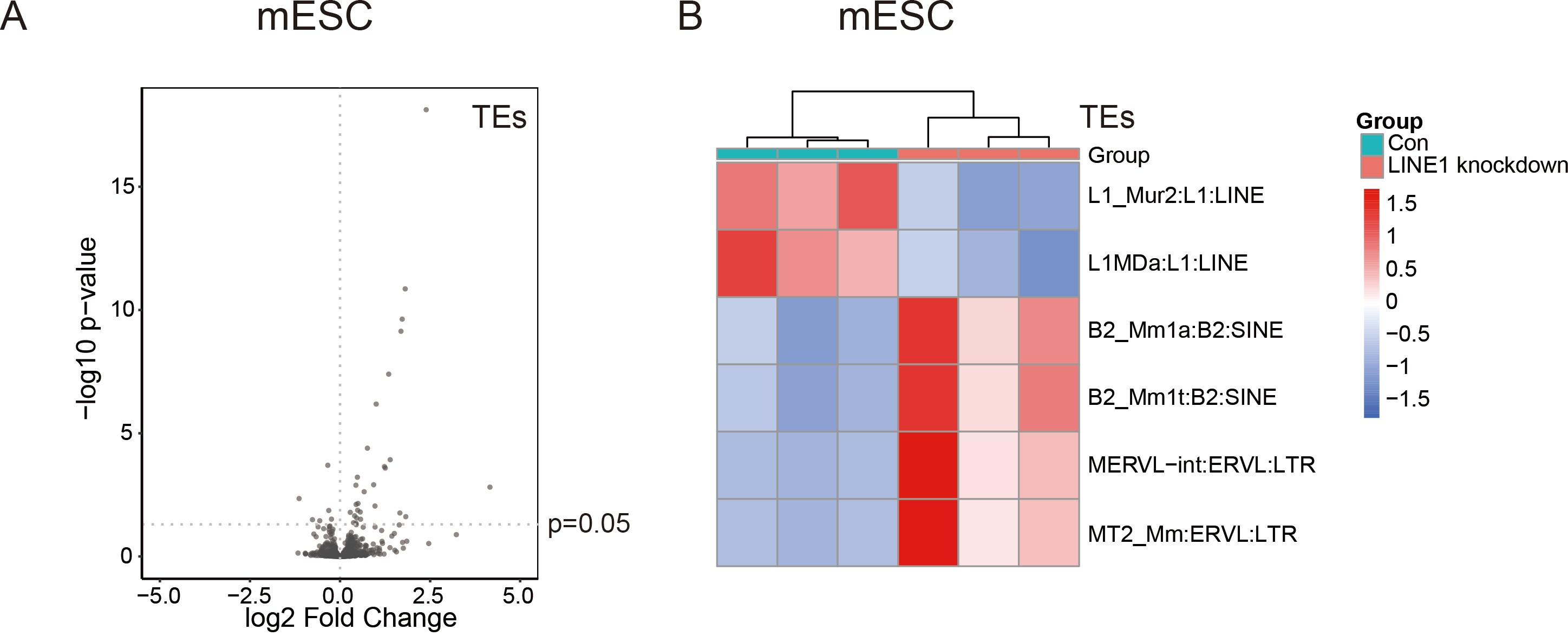

**Fig. S4.** Potential crosstalk between different TE subfamilies.

(A) Knockdown of LINE1 in mouse embryonic stem cells (mESCs) led to differentially expressed subfamilies of TEs.
(B) Heatmap shows top 2 misregulated LINE, SINE and LTR subfamilies upon LINE1 knockdown in mESCs.

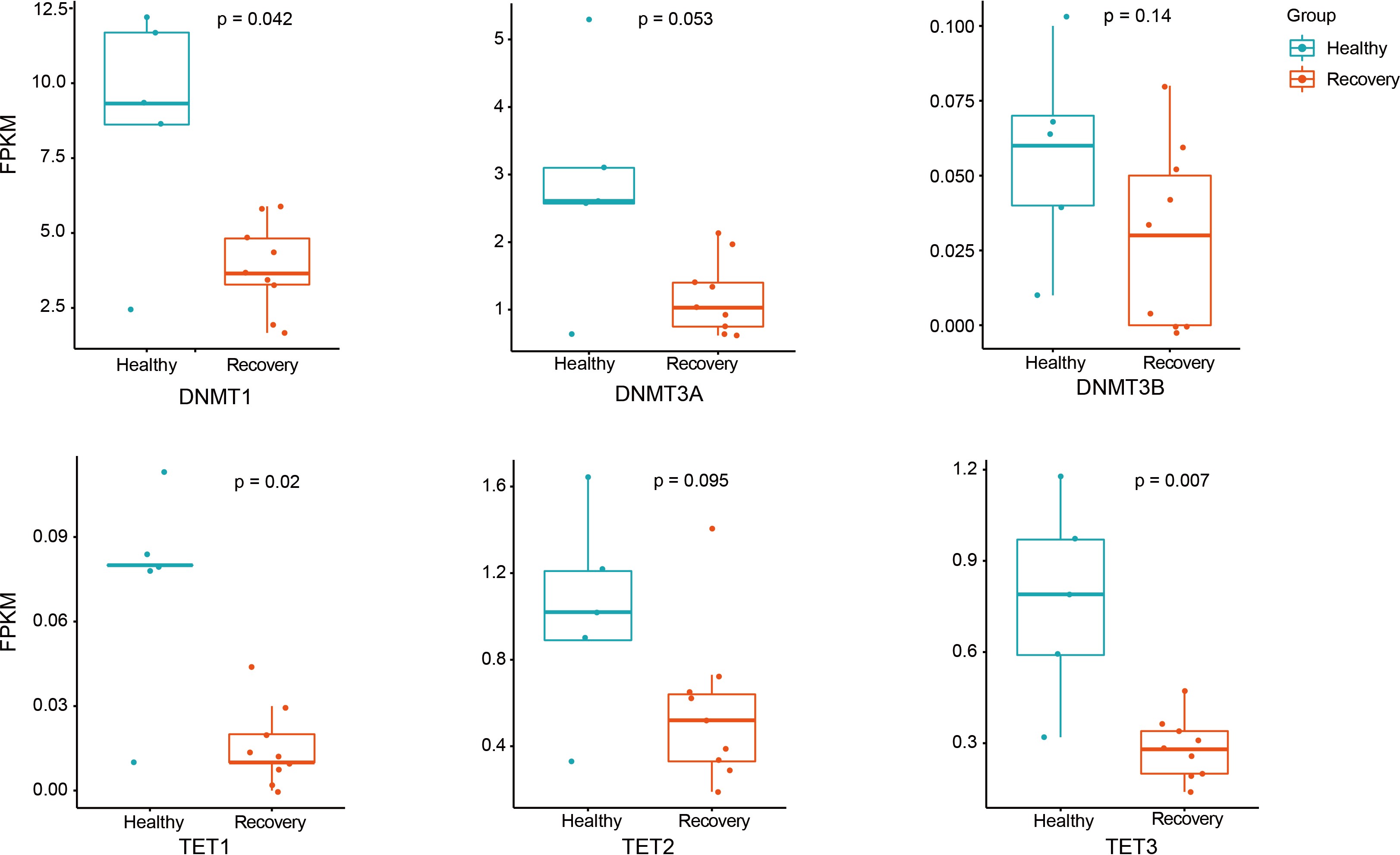

**Fig. S5.** Boxplot displaying downregulation of genes regulating DNA methylation. DNMT1, DNMT3A, DNMT3B, TET1, TET2, TET3 are shown, and each dot represents one sample. The median, first, and third quartiles are shown. Two-sided Wilcoxon signed-rank test were used for the comparisons.
